# Multi-faceted Semantic Clustering With Text-derived Phenotypes

**DOI:** 10.1101/2021.05.26.21257830

**Authors:** Luke T Slater, John A Williams, Andreas Karwath, Hilary Fanning, Simon Ball, Paul Schofield, Robert Hoehndorf, Georgios V Gkoutos

## Abstract

Identification of ontology concepts in clinical narrative text enables the creation of phenotype profiles that can be associated with clinical entities, such as patients or drugs. Constructing patient phenotype profiles using formal ontologies enables their analysis via semantic similarity, in turn enabling the use of background knowledge in clustering or classification analyses. However, traditional semantic similarity approaches collapse complex relationships between patient phenotypes into a unitary similarity scores for each pair of patients. Moreover, single scores may be based only on matching terms with the greatest information content (IC), ignoring other dimensions of patient similarity. This process necessarily leads to a loss of information in the resulting representation of patient similarity, and is especially apparent when using very large text-derived and highly multi-morbid phenotype profiles. Moreover, it renders finding a biological explanation for similarity very difficult; the black box problem. In this article, we explore the generation of multiple semantic similarity scores for patients based on different facets of their phenotypic manifestation, which we define through different sub-graphs in the Human Phenotype Ontology. We further present a new methodology for deriving sets of qualitative class descriptions for groups of entities described by ontology terms. Leveraging this strategy to obtain meaningful explanations for our semantic clusters alongside other evaluation techniques, we show that semantic clustering with ontology-derived facets enables the representation, and thus identification of, clinically relevant phenotype relationships not easily recoverable using overall clustering alone. In this way, we demonstrate the potential of faceted semantic clustering for gaining a deeper and more nuanced understanding of text-derived patient phenotypes.

## Introduction

In clinical settings, text narrative is the primary source of record, and thus the most detailed and complete source of phenotypic information concerning patients. As such, natural language processing provides an important set of tools for information extraction, which can lead to improved insight into biomedical and clinical entities [1].

Natural language processing challenges can be reduced to the problem of resolving ambiguity [2]. Ontologies aid the resolution of ambiguity through their provision of logical features, controlled domain vocabularies, and data linkage [3]. Thus, biomedical ontology and text mining are deeply interlinked. Ontologies are standard tools in text mining, frequently used to construct vocabularies for named entity recognition (NER) and subsequent entity linking in text, thereby also enabling secondary analyses of linked concepts that characterise biomedical text [4].

Semantic similarity has become an established method for the exploitation of clinical text to determine the interrelationships between patients, diseases and phenotypes. [5]. Semantic similarity methods have long been used in semantic analysis of human language, to explore relatedness between words and similar constructs. Later, these concepts were explored in the context of biomedical ontology and have been used with success for predicting protein-protein interaction [6] and measuring functional similarity between genes [7].

Semantic similarity has been applied for use in differential diagnosis of rare diseases, particularly using the Human Phenotype Ontology (HPO) [8]. This has typically been performed through use of semantic similarity to compare expertly curated patient phenotype profiles with reference databases that define phenotypes typically associated with particular rare diseases [9], such as OMIM [10].

We have previously explored the use of text-derived phenotype profiles for differential diagnosis of common diseases, comparing patient-patient comparisons and patient-reference similarity, and demonstrating a superior method in a hybrid approach that extended literature-derived disease profiles with in-context information mined from clinical text [11]. Several other studies have investigated the use of text-derived data to build and extend phenotype profiles based on semantic similarity, such as Doc2HPO [12], which explored both uncurated and curated text-derived phenotypes for use in rare disease diagnosis and gene variant prioritisation, identifying a hybrid approach exhibiting the best performance. Other studies has focused on using text data to extend the background knowledge used in these methods [13, 14]. Methods for differential diagnosis and patient stratification using free text that do not rely on semantic similarity, using a range of statistical and machine learning methods, are also common [15].

Semantic similarity has also been applied to clustering tasks. Semantic similarity, in the context of performing a pairwise comparison between all entities, results in a similarity matrix, which can easily be transformed into a distance matrix used for clustering. Clustering based on semantic distance matrices has been used in biomedical literature curation [16–18] but has also formed the basis for associating genetic mutations to patient phenotype profiles [19]. Phenotype profiles have been used, with and without ontological annotation, to discover subtypes of autism, Alzheimer’s, and schizophenia [20–24]. In contrast to this work, underlying patient symptoms in these studies were derived largely from diagnostic tools. Semantic similarity has also been used in co-clustering tasks and in the evaluation of clusters generated from measured biological data. The Gene Ontology has served as a basis for explaining the functionality of clustered genes and protein arrays [25–27]. Recent work has derived symptom clusters from OMIM [10] and other databases, and validated those clusters by investigating genomic profiles associated with them [28]. These comparisons can be thought of as multi-view approaches, where one form (view) of data is used to assess the stability of clusters derived from another view. Outside of these situations, graphical, and statistical approaches have been suggested [29–31].

While semantic similarity is undoubtedly a powerful tool, it is limited by virtue of collapsing similarity between entities characterised by complex sets of ontology terms a single score. This both loses and obscures information by which entities can be compared, and makes it more difficult to extract meaningful relationships from the resulting similarity matrix.

In this article, we first explore and explain limitations to the use of singular similarity scores for comparing patient phenotype profiles. Based on this understanding, we propose a solution to these problems by calculating multiple semantic similarity scores for each profile, based on different subsets of their annotations, derived from different subsets of the source ontology (facets). We demonstrate this approach by creating text-derived phenotypic annotations for patients from MIMIC-III, and splitting them into separate facets, revealing that annotations are widely spread across multiple different facets of HPO. We then calculate the semantic similarity between patient visits for each facet, and then generate clusters for each of the facets, as well as for all annotations. We also present a new method of identifying qualitative explanatory variables for semantic clusters, and use this method to evaluate and explore the different sets of clusters. We show that facet-based clustering provides the opportunity to identify different modes, features, and aspects of entity similarity. We then demonstrate that consideration of multiple faceted semantic similarity matrices can lead to greater insight into datasets characterised by text-derived phenotype profiles, opening up the potential for future development of a method that combines the additional information with less loss of information than a single semantic similarity score.

### Limitations of Singular Semantic Similarity

In this section, we will explore two popular combinations of similarity score, and show how they may lose information or obscure relationships between entities. In particular, we will explore Resnik pairwise similarity [32], max and Best Match Average (BMA) groupwise similarity [33], and Resnik corpus-based information content [32]. Figure 1 shows the equations by which these measures can be calculated.

**Figure 1.**
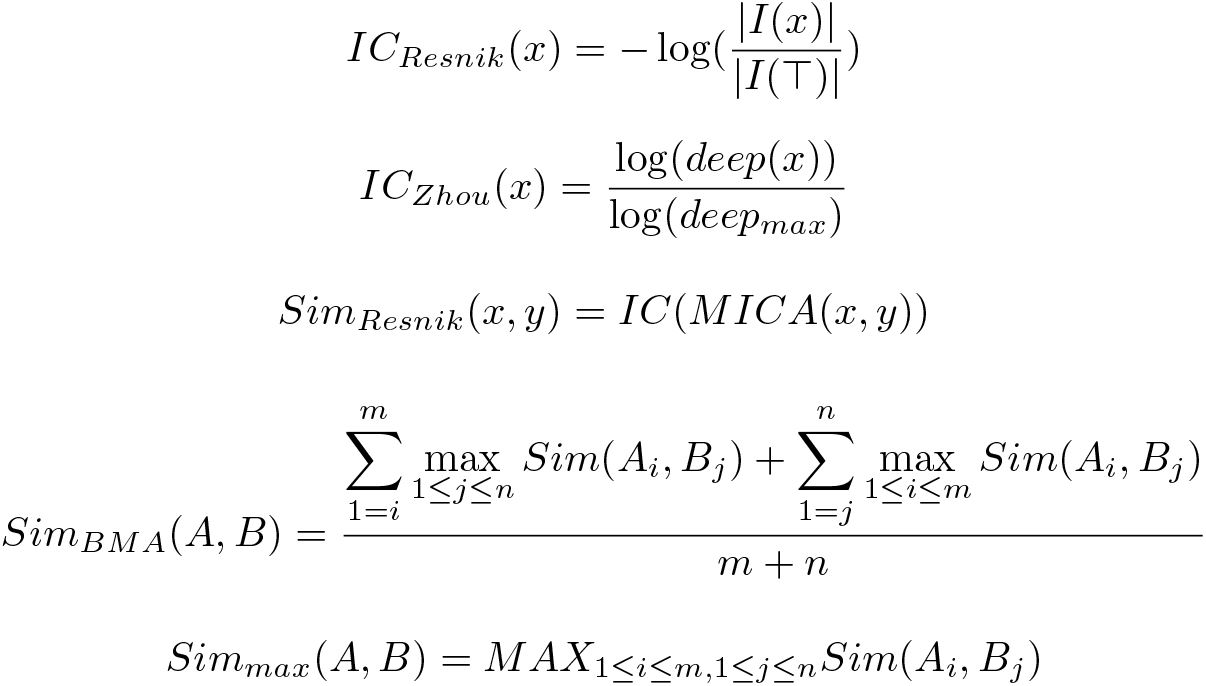
Equations for Resnik corpus-derived and Zhou structural information content measures, Resnik pairwise similarity, and Best Match Average and max indirect groupwise similarity. Groupwise similarity measures accept two sets of ontology terms, while pairwise similarity measures accept two ontology terms.

The max groupwise method identifies the greatest similarity score between members in two sets of terms, and employs it to characterise the overall similarity between the two sets. Meanwhile, BMA recovers, for each term in each set, the pair with the greatest similarity score, and then averages them, to produce the final similarity score between the sets. BMA can alternatively be thought of as generating a pairwise similarity matrix between all single terms in both sets, then selecting the maximal value for each row and column, and then averaging them. These groupwise measures are indirect, requiring a pairwise similarity measure to determine the individual term similarity, which is then used to calculate a final score. In this case, the Resnik measure of pairwise similarity is employed, which is defined by the information content of the most informative common ancestor of the two classes; that is the ontology superclass that subsumes both classes, and has the greatest information content. Information content is then represented by the negative log probability of that class appearing in a corpus (often the set of profiles being considered for comparison).

Figure 2 shows three patients whose phenotypes are described using classes from the Human Phenotype Ontology (HP), as sets of terms in the ontology. All patients share `abnormal heart valve morphology (HP:0001654)`, while only *P*_2_ and *P*_3_ share `increased inflammatory response (HP:0012649)`. In the calculation of *Sim*_*max*_(*P*_2_, *P*_3_), if `abnormal heart valve morphology (HP:0001654)` has a greater information content than `increased inflammatory response (HP:0012649`), then (*P*_2_, *P*_3_) will have the same semantic similarity score as *Sim*_*max*_(*P*_1_, *P*_2_), and *Sim*_*max*_(*P*_1_, *P*_3_), despite them sharing an additional inflammatory phenotype that neither share with *P*_1_. This is mitigated somewhat by the corpus-derived information content: if these were our only three patients, then *Sim*_*max*_(*P*_2_, *P*_3_) would likely instead find the maximal match in `increased inflammatory response (HP:0012649)`, since this is the ‘rarer’ phenotype. However, many semantic similarity configurations do not use annotation frequency-derived measures of information content. Nor do such configurations present a reliable solution, since the estimation of IC could easily be made in the context of a corpus that rarely discusses cardiac phenotypes, for example in a population of patients with behavioural disorders. Furthermore, even in the case that *Sim*_*max*_(*P*_2_, *P*_3_) does select the inflammatory phenotype, the similarity of their cardiac phenotype is then only encoded in their similarity to *P*_1_. This kind of indirect relationship is difficult to recover when exploring or making secondary use of the similarity matrix. Furthermore, it requires that there be other patients whose maximal match with *P*_2_ and *P*_3_ is found via that particular secondary phenotype. If *P*_1_ did not exist, did not have a cardiac phenotype, or had another non-cardiac phenotype that better matched *P*_2_ or *P*_3_ (such as, in a minimal example, an inflammatory phenotype), there would be no record of the cardiac similarity between *P*_2_ and *P*_3_ in the resulting similarity matrix.

**Figure 2.**
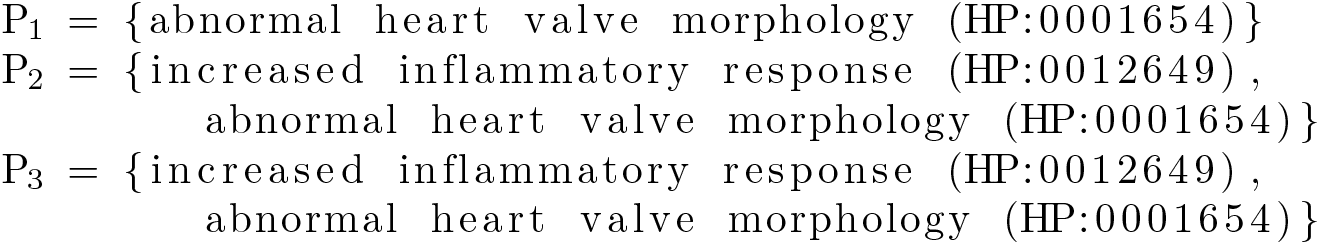
Three exemplary patient phenotype profiles. A phenotype profile for a patient can be characterised as a set of ontology terms associated with the patient. In this case, the patients are characterised by terms from the Human Phenotype Ontology.

BMA was designed to ameliorate these problems, by allowing the influence of every term in each set in the final similarity score. In the calculation of *Sim*_*BMA*_(*P*_2_, *P*_3_), `increased inflammatory response (HP:0012649)` in *P*_1_ would be compared to both `increased inflammatory response (HP:0012649)` and `abnormal heart valve morphology (HP:0001654)` in *P*_2_, finding its ‘best match’ in `increased inflammatory response (HP:0012649)`. The same would be repeated for `abnormal heart valve morphology (HP:0001654)` in *P*_1_, finding its best match in the correlated cardiac phenotype. The same calculation would be performed in reverse between *P*_2_ and *P*_1_. The final similarity score would be the average of the four resulting scores, thus encoding both the similarity of their inflammatory and their cardiac phenotypes. However, this can still easily lead to confounding results. If, for example, the pairwise value of the comparison between the inflammatory terms is lower than that between the cardiac terms, depending on the similarity between the cardiac and inflammatory terms, the result could be unexpected. For example, in Figure 3, we define the values for *Sim*_*Resnik*_ between the three HPO terms under consideration. Then, we evaluate *Sim*_*BMA*_(*P*_1_, *P*_2_) and *Sim*_*BMA*_(*P*_2_, *P*_3_), finding that the similarity between *P*_1_ and *P*_2_ would be greater than the one between *P*_2_ and *P*_3_, despite the latter sharing an additional phenotype, and despite the individual pairwise values correctly ‘ranking’ the similarities between them. In reality, it’s likely that IC values would be much less clear, especially with an annotation frequency derived from text rather than manually curated databases, enhancing this effect. Furthermore, when the correct best matches are found in BMA, similarity is nevertheless encoded into a singular value via averaging the best matches for each constituent term, thus indirectly encoding those individual dimensions with loss of information. In addition, the singular scores obscure multiple similarity and make it difficult to extrapolate relationships, in the same way as it does for max.

**Figure 3.**
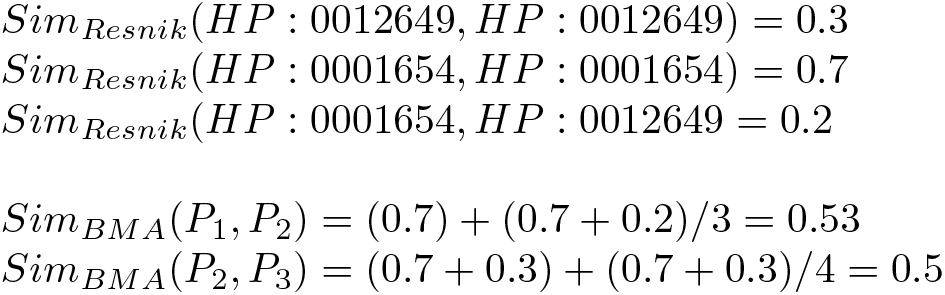
Example values for pairwise similarity between the terms contained in the example corpus, and associated BMA calculations for two pairs of example patients (per Figure 2). In the example, the two patients that share a cardiac and an inflammatory phenotype are judged as less similar than two patients who only share a cardiac phenotype, showing how BMA can easily lead to confounding results.

These issues are compounded as the set of phenotype profiles becomes larger and more complicated. They are also apparent across other semantic similarity and information content measures. Structural IC measures, such as the Zhou method (also shown in Figure 1), instead employ measures of depth or distance in an ontology graph to determine information content [34]. In addition to losing additional information from secondary qualities, such approaches may also not match on the most ‘important’ feature for a particular context. Since ontologies reflect current trends in scientific interest, their development, complexity, and structure across different facets correspond to the domains reflecting these trends and are not necessarily aligned with the complexity, structure, or importance of the conceptual area itself [35]. This is a source of bias, and may be a particular limitation for data-led approaches that intend to infer new knowledge or propose new hypotheses.

Across real-world datasets, especially in relation to text-derived data, phenotype profiles are likely to contain many phenotypes that describe different and disparate features of a patient’s condition. Particularly in clinical environments, we hypothesise that multi-morbidity and complex diseases will lead to representation of phenotype annotations across organisational and systemic phenotype structures for single admissions. By focusing only on a maximal, or averaged ‘best match’ phenotype comparison, additional features that can form the basis on which patients can be contrasted are unlikely to be identified.

Moreover, a recent study, examining how annotation size affects semantic similarity calculations, demonstrated that entities with many annotations exhibit increased similarity across them, even in the absence of any biological similarity [36]. This is especially relevant to text-derived complex phenotypes, which can potentially encompass many phenotype classes. In a previous study, 1,380,216 HPO annotations were associated with clinical narratives related to 1,000 patient visits reported in MIMIC-III, with an average of 1380.2 annotations per patient visit [37]. In contrast, another study, examining clustering based on HPO phenotype profiles, manually curated by experts, reported an average of 7.5 annotations per case [38]. This effect is corroborated by work using Doc2HPO, which demonstrated a vast improvement in performance when using manually curated text-derived phenotype profiles to diagnose rare genetic diseases (however, this effect cannot easily be separated from the improvement in the quality of annotations, which was the primary objective of the curation process) [12]. Thus, unmitigated use of text-derived phenotypes for semantic similarity may incur unintended bias under analysis, on account of the large phenotype profiles they produce. This may be particularly pronounced for clustering approaches, which may not perform well in an environment in which all entities are very similar, since fewer differences between groups of entities can be discerned.

## Methods

The software used to run these experiments is freely available, including documentation, from https://github.com/reality/facetsim. All implementation was performed with the R programming language version 3.6.3 [39], and Groovy version 2.4.16 [40].

### Data Preparation and Information Extraction

We sampled 1,000 patient admissions (clinical encounters) and their associated narrative text from MIMIC-III [41]. MIMIC is a freely available database describing nearly 60,000 admissions in a critical care setting at Beth Israel Deaconess Medical Center in Boston, Massachusetts, and includes clinical text amongst a wealth of structured data concerning the patient admissions/clinical encounters [41]. Most encounters or admissions to ICU are single events for a specific individual but a small number represent repeat admissions over an 11 year period (22.4%). Our sample of 1,000 admissions described 982 unique patients, with each of the 1,000 admissions being treated separately in our analysis. The median age of adult patients is 65.8 years and the median length of time spent in ICU is 2.1 days, meaning that the data in MIMIC-III is largely concerned with acute onset or critical disease, or immediate post-interventional support. Admissions are coded for diagnoses that were produced by professional clinical coders, using the ICD-9 terminology. Sampled admissions were limited to those with diagnoses mapped to the Disease Ontology (DO), since we wished to retain the sampled admissions, annotations, and associated clusters for further secondary analysis using DO.

The clinical narrative for patient visits is stored in different note events, which occur at different points of time during the visit. We concatenated all note events for each patient visit into a single file. We further applied pre-processing measures to remove extra whitespaces, collapsing multiple line breaks into sentence breaks. We then used the Komenti semantic text mining framework [42] to extract HPO terms from the text. HPO is a biomedical ontology that describes abnormal observable properties of humans [8]. This produces a list of concepts associated with each patient admission, by virtue of a label associated with that concept appearing in the text associated with that patient visit. These annotations are then used to form a patient phenotype profile, which is a set of HPO phenotypes associated with that patient’s encounter.

### Facets

To determine the HPO facets, we used Komenti to query HPO for direct subclasses of `Phenotypic abnormality (HP:0000118)`, which gave us a list of 20 high-level phenotype categories in the ontology. We then performed transitive subclass queries on each of those classes, to identify the full list of classes that belong to each. These 20 high level classes, and their subclasses, form our facets. To facilitate facet-based analysis of our data, we created separate phenotype profiles for each patient, one for each facet, and containing only the annotations that belong to that facet.

### Semantic Similarity

In order to compare facet-derived patient-patient similarity with that using the whole ontology, we first calculated a similarity matrix describing pairwise similarity between all patient visit phenotype profiles, using the full set of HPO annotations associated with each patient visit. We then created one additional similarity matrix for each facet, using the phenotype profile subsets containing only annotations belonging to the particular facet.

We used Resnik pairwise [43] similarity with Best Match Average [33], with the Resnik method of information content. The equations for these measures are shown in Figure 1. Finally, we calculated the values using the Semantic Measures Library (SML) [44].

### Clustering

So as to generate the similarity matrices for clustering, we first converted them into adjacency network matrices, using a soft thresholding setting of 2 (raising the distance function result to the power of 2) to minimise the effect of small differences between similarity scores [45]. We then converted the adjacency matrix into a topological overlap matrix [46], and subtracted it from 1 to provide the final distance matrix used for clustering. Matrix pre-processing steps were performed using the WGCNA R package [45].

Kmeans was used to cluster the resulting distance matrix. Optimisation of cluster centers was performed automatically, selecting *k* clusters with *k* ∈ {3, …, 20} with the maximum silhouette score. 2 clusters were excluded from consideration, because facet clusters would always select this number, on account of the split between patients with and without any annotations in that facet. We evaluated the resulting clusters using the silhouette score, as well as visual investigation.

Visualisations were produced using the factoextra R package [47], which plots clustering partitions using principal components of the data.

In addition, we developed an algorithm for determining qualitative explanations for semantic clusters, described in the results. The algorithm identifies ontology terms that more exclusively identify the patient phenotype profiles in the given cluster. We applied this to our overall clusters to identify top-level groups of patient phenotypes. We then examined a particular example of neoplasm, applying the explanation algorithm to the neoplasm clusters, to find neoplasm phenotypes that particularly characterised those clusters.

To further characterise the neoplasm clusters, we explored an alternative application of the explanation algorithm. We continued to explain the neoplasm clusters, but use candidate explanatory terms from a *different* facet. We hypothesised that by doing this we would be able to recover relationships between particular neoplasm phenotypes and non-neoplastic phenotypes.

## Results

### Text-derived Phenotypes and Facets

We sampled 1,000 patient admissions, describing 982 unique patients. We annotated each admission with HPO terms using Komenti, as described in the methods. This yielded 43,953 annotations, which were then sorted into their facet groups (transitive subclasses of the direct subclasses of `Phenotypic abnormality (HP:0000118)` in HPO). Table 1 shows the number of annotations and the number of admissions in the sample with at least one annotation for that facet. Annotations are distributed throughout the facets, with counts much smaller than the overall group. The largest facet group, `abnormality of the cardiovascular system` with 10,057 annotations, remains nearly four times smaller than the total number of annotations for all facets. This is consistent with the overall composition of the MIMIC-II population where ICD:414.01 (‘Coronary atherosclerosis of native coronary artery’) and ICD: 410.71 (‘Subendocardial infarction, initial episode of care’) are together the most common ICD-9 annotations in the complete MIMIC dataset, and make up 10.7% of admission annotations.

**Table 1.**
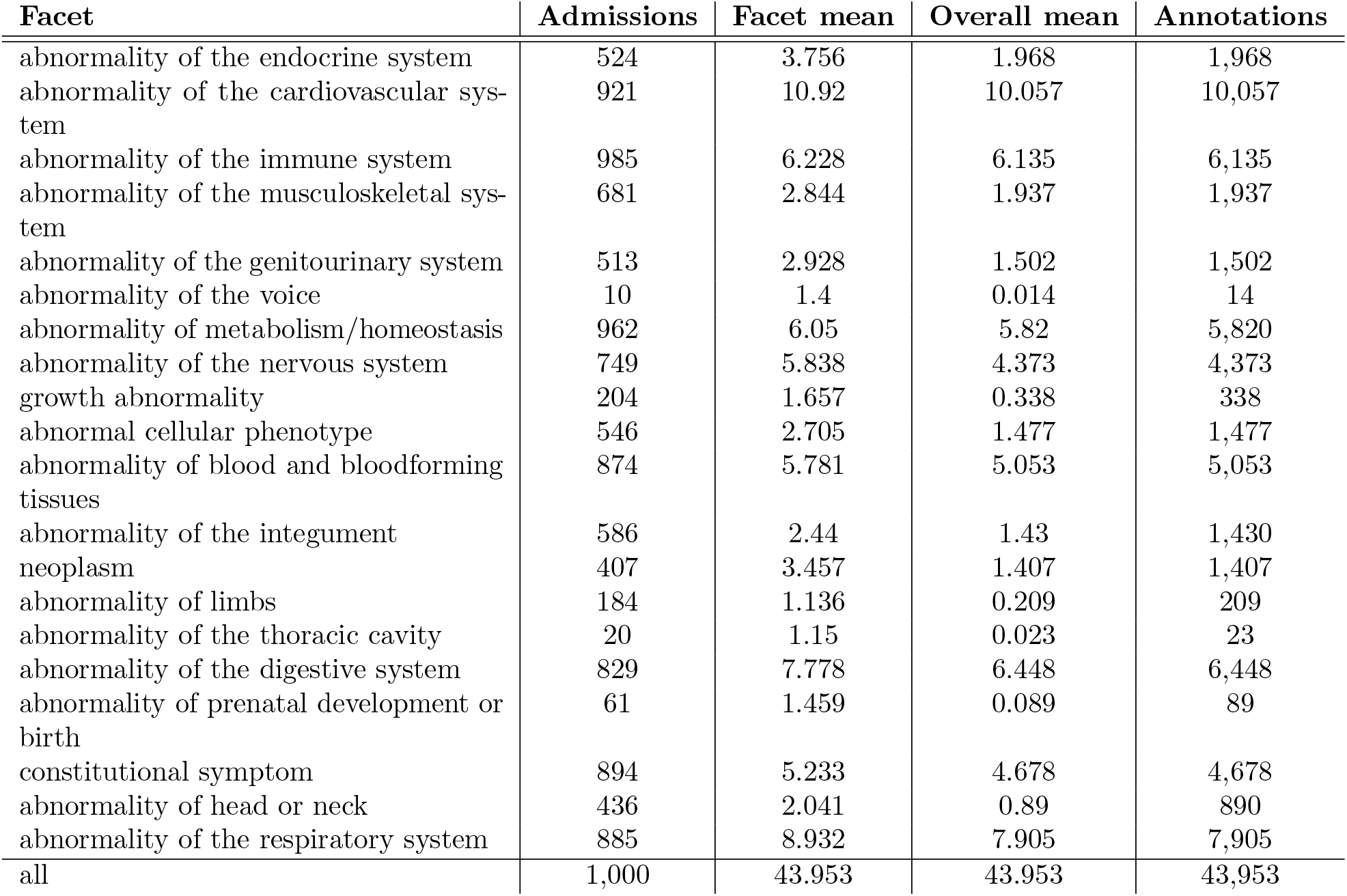
Text-derived patient phenotype per-facet. Concept annotations were sorted into facets according to which subclass of `Phenotypic abnormality (HP:0000118)` they fall under, which in some cases may be several. Admissions refers to the number of admissions out of the sample of 1,000 that had at least one annotation in that facet. Facet mean refers to the mean number of annotations per patient only for the patients that had at least one annotation in that facet, while overall mean is the mean number of annotations per patient overall (out of the total 1,000 patient admissions sampled).

The mean number of overall annotations for facets in our sample of 1000 admissions is 3.09, in other words the average number of annotations involved in any semantic similarity comparison within a single facet is 3.09. This results in a reduction of more than a factor of ten compared with the average of 43.953 for all facets considered together.

There is also a large range of 10,043 in the number of annotations belonging to the facets, as well as a range of 911 for the number of admissions described. In both cases, these figures represent ranges between maximal and minimal possible values for these variables. As such, there are several facets with very few annotations, particularly `abnormality of the voice, abnormality of the thoracic cavity`, and `abnormality of prenatal development or birth`, which all have fewer then 100 annotations, and describe fewer than 100 patients. Meanwhile, abnormality of the cardiovascular system, `abnormality of the immune system`, and `abnormality of metabolism/homeostasis` describe more than 90% of patients. This reflects the overall composition of the MIMIC-III population as characterised by ICD-9 annotation. The cardiac care and coronary recovery units together account for 33.1% of admissions and the medical intensive care unit 39.5%. The HPO class `abnormality of the thoracic cavity` does not contain any phenotypes of the cardiovascular system or heart and therefore there are few annotations to this facet, despite the heart being contained within the thoracic cavity. Meanwhile, few admissions and annotations were associated with the `abnormality of prenatal development or birth`, despite MIMIC-III describing 7,800 neonatal admissions. This can be explained by our sampling method, which was limited to patients with primary diagnoses that were mapped to DO. The mapping contains only 13 neonatal codes (ICD-9 range 630-679), which would limit the appearance of neonatal annotations due to the exclusion of neonatal admissions without mappings to DO.

13 facets describe more than 50% of admissions, showing that there is coverage of the same admissions by several facets. This is confirmed by considering how many facets contain classes to which a patient is annotated. The mean number of facets involved in patient annotation is 11.271. With a few exceptions noted above, most facets are well-represented across the patient phenotype, and thus splitting annotations into these groups provides phenotype profiles describing different features of the patient phenotype.

### Cluster Scoring and Explanation Algorithm

In finding explanatory terms for clusters or groupings derived from sets similarity of ontology classes, we want to optimise three conditions:

### Inclusivity

Terms that more members of the cluster share.

### Exclusivity

Terms that more exclusively identify the cluster; fewer members of other clusters share these terms.

### Specificity

Terms that are more specific; more informative; less generic.

Our approach uses the definitions of three scores that measure these properties of candidate explanatory classes. In addition, in order to optimise inclusivity, we can leverage the subsumptive hierarchy of the source ontology from which the semantic similarity measurements were derived. For example, assumming a class A, with subclasses B and C, then A may be used as an explanatory term that includes both B and C. To do so, we generate scores for all terms present in the cluster, as well as all of their superclasses.

The inclusivity score is defined by the number of members of a cluster that contain a transitive subclass of the given class. The exclusivity score is defined by the the number of patients in every other cluster that contain a transitive subclass of the given class, and is subtracted from one, so that greater values reflect greater exclusivity. The specificity score is defined as the information content of the class. These can be formally defined as shown in Figure 4, where O is the set of classes in the ontology, and C is the set of clusters. Each element represents each member of that cluster, which is in turn a set that contains the list of ontology terms associated with that member.

**Figure 4.**
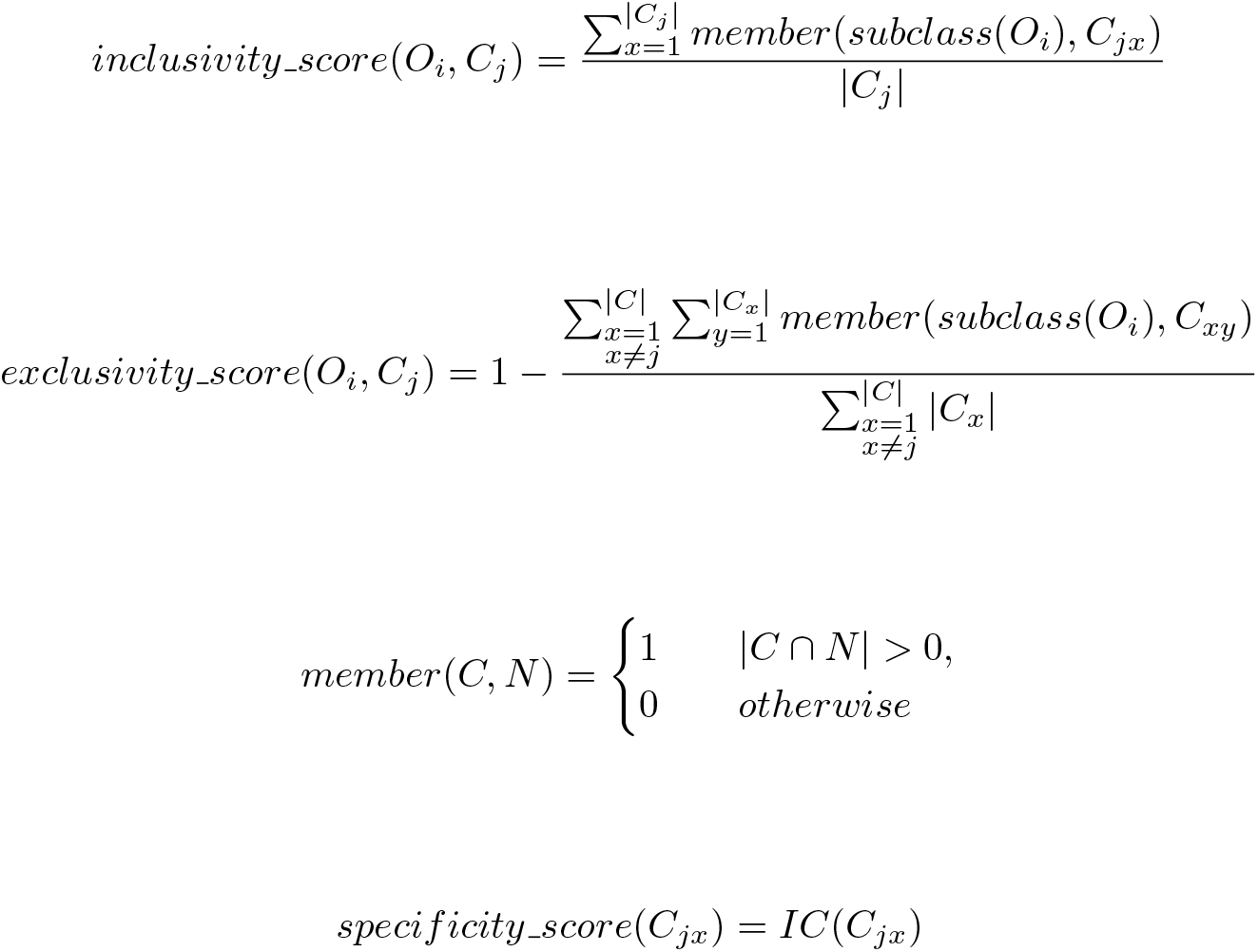
Equations for inclusivity, exclusivity, and specificity scores, which are used to evaluate how well ontology terms characterise a cluster. O is the set of classes in the ontology, and C is the set of clusters. Each element of C represents a cluster, as a set of cluster members that contains the set of ontology terms associated with that member. The subclass function returns a set containing the transitive subclasses of the class given in the parameter. These scores are calculated for all classes in the ontology, which are considered candidate explanatory terms passed into Algorithm 1.

Together, these scores measure, for each combination of class and cluster, how much it explains the cluster, how little it explains the other clusters, and how specific the term is. In order to recover high quality explanatory variables, terms with optimal combinations of these values need to be identified. So as to identify sets of high quality explanatory variables for clusters, the set of explanatory variables needs to explain a large number of the members of the cluster. To measure this, the overall inclusivity score is introduced, defined by the percentage of the cluster membership covered by at least one explanatory variable in the set.

To optimise these variables, we developed an algorithm that steps down through acceptable values of the above defined heuristics until a suitable set of explanatory variables is identified. Algorithm 1 represents the developed algorithm, while descriptions of the parameters and hyperparameters are described in Table 2. Maximal and minimal values are given as parameters for the information content, the exclusion score, and the inclusion score, while the total inclusivity score is only assigned a maximum value. Cutoffs for the relevant scores are derived from these parameters, which are stepped down until a satisfactory set of explanatory terms are found, where both the satisfaction and the order of cutoff is defined by the algorithm’s order of variable importance:

**Table 2.** List of parameters used in Algorithm 1. The hyperparameter settings used in this article, except where otherwise stated, are given in Table 9.

| Variable | Description |
| --- | --- |
| MAX_IC | Maximal acceptable value for information content. |
| MIN_IC | Minimal acceptable value for information content. |
| MAX_INCLUSION | Maximal acceptable value for inclusion score. |
| MIN_INCLUSION | Minimal acceptable value for inclusion score. |
| MAX_EXCLUSION | Maximal acceptable value for exclusion score. |
| MIN_EXCLUSION | Minimal acceptable value for exclusion score. |
| MAX_TOTAL_INCLUSION | Maximal acceptable value for total inclusion coverage. |
| STEP | The step by which to reduce heuristics each iteration. |
| EXPLANATIONS | Full unordered set of explanations for the cluster. |
| FACET | Explanatory terms will be limited to terms in the given facet. |

1. Total inclusion score
2. Specificity/Information content
3. Inclusion and exclusion

This order of preference renders that inclusion and exclusion are first stepped down to their minimal values (simultaneously and symmetrically), before information content is stepped down, followed by the total inclusion score when information content minimum cutoff is reached. At each step-down, all cut-offs with a lower priority are reset to their maximal values defined by the relevant parameter. Values for the algorithm parameters used in this paper (except where otherwise noted) are given in the appendix (Table 9).

Since the algorithm involves determining a satisfactory solution depending upon a number of thresholds and boundaries, the algorithm can be described in relation to a Multi-Objective Optimization Problem (MOOP). In this task, our ‘optimal’ solution is a set of explanations that maximises the heuristics, such as inclusivity score and IC, which can in this expression be termed as objective functions. The solution given above can be expressed as a variation of the *ε*-constraints method, in which one objective function is retained, while the others are expressed as a series of constraints [48]. In this case, the *total inclusion score* can be considered the objective function, as it is the heuristic with the highest priority, while all others are constrained functions whose limits are defined by the user-provided parameters (described in Table 2). The variation from the *ε*-constraints method consists in the stepwise reduction of maximal constraint values in the order of the parameter priority given above, until a solution is found that maximises the objective function (*total inclusion score*) within the boundaries of the maximal/minimal settings of the constrained parameters and their priorities. Though this is expressed in the above description as ‘stepping down’ through values of *total inclusion score* until a solution is found, this is equivalent to its maximisation as an objective function.

#### Algorithm 1

Algorithm for identifying characterising ontology terms for a cluster, by stepping down through exclusion and inclusion scores.

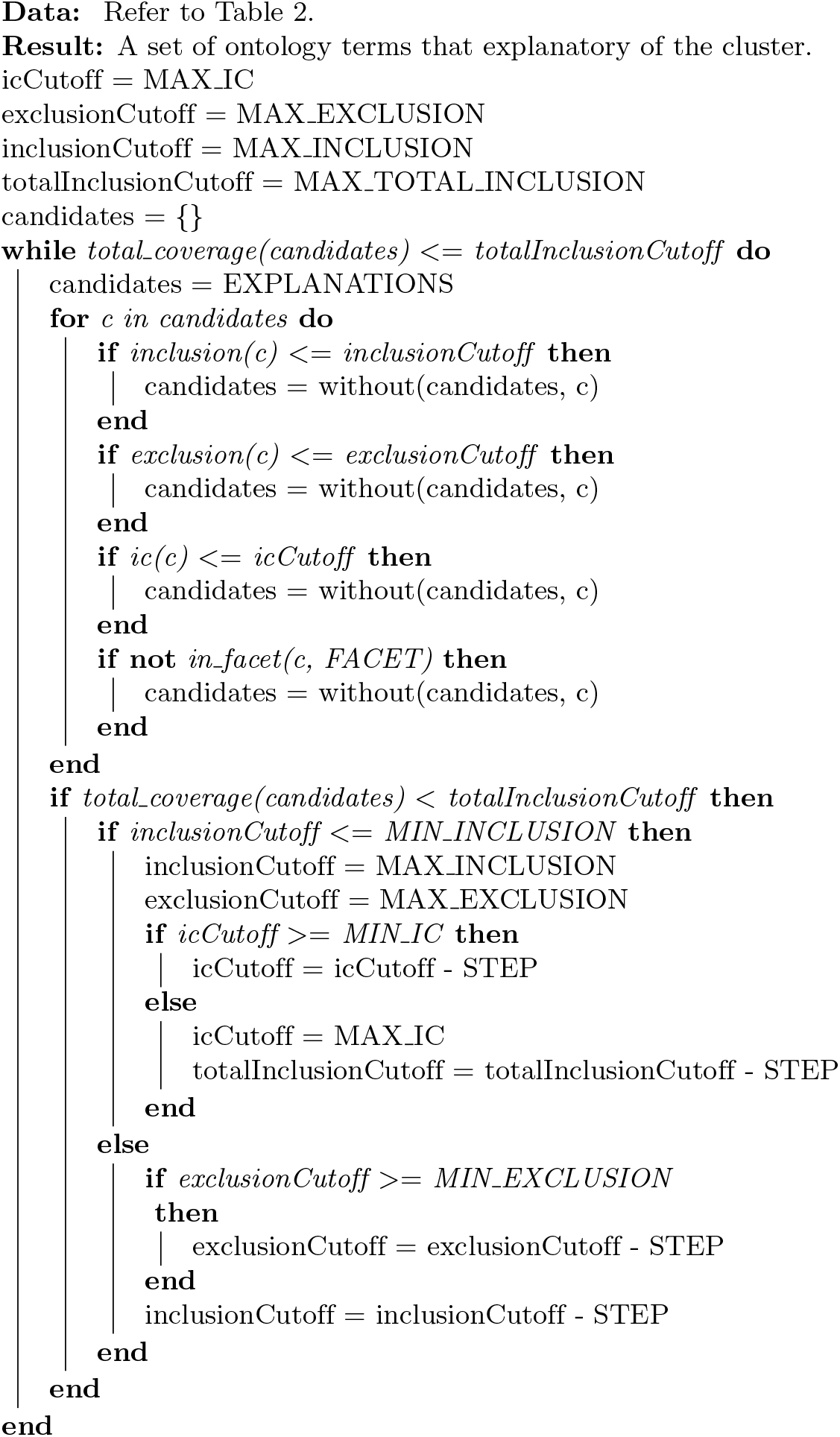

### Overall Clustering

We first generated clusters based on the semantic similarity matrix for all annotations associated with our patient sample. The largest silhouette score was 0.215 with three clusters, which indicates a relatively poor representation of the underlying structure. Figure 5 shows a visualisation of the clusters.

**Figure 5.**
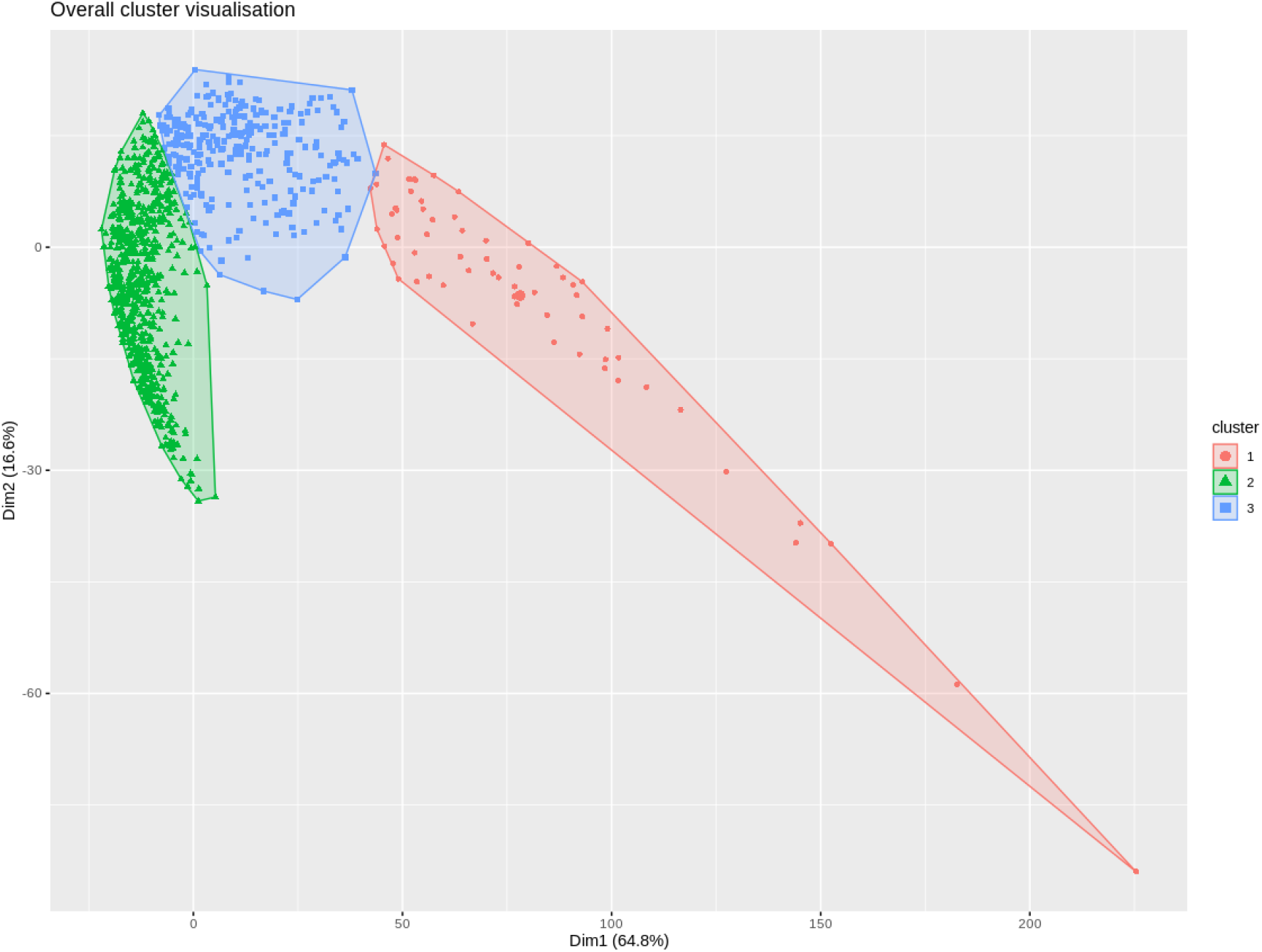
Visualisation for overall patient visit clustering on the basis of text-derived HPO phenotype profiles. Cluster 1 contained 63 patients, while cluster 2 contained 653 patients, and cluster 3 contained 284 patients. Explanations derived for the clusters can be seen in Table 3.

We then ran our cluster explanation algorithm on the clusters, receiving qualitative HP explanations for each cluster, which can be seen in Table 3. Despite the low silhouette score, cluster explanations, with large overall inclusivity, were found for all clusters, with an average value of 89.6% coverage of cluster members. However, for individual explanatory terms, the average inclusion and exclusion scores were 42% and 55% respectively. This indicates that fewer terms with high individual explanatory power for clusters were found. The term with the greatest inclusion score was `abnormal cellular phenotype (HP:0025354)` in cluster 2. Given, however, that this term also appeared in explanatory sets for clusters 1 and 3, with similar inclusion ratios, indicates that this does not uniquely distinguish any of the clusters. There were a number of higher exclusion scores, such as `abnormal heart valve morphology (HP:0001654)` and `abnormal aortic valve physiology (HP:0031652)`, which have exclusivity scores of 0.68 and 0.66, despite the cluster only accounting for 284 total patients. The contrast of high overall inclusivity scores with the lower inclusivity and exclusivity scores of individual terms reveals that these patient phenotypes are highly multi-morbid, rather than being characterised and groupable by individual conditions or phenotypes.

**Table 3.** Explanations for overall clusters

|  |  |  |  |
| --- | --- | --- | --- |
| **Cluster 1 (63 patient visits, overall inclusivity: 77.78)** | **Exclusion** | **Inclusion** | **IC** |
| abnormal cellular phenotype (HP:0025354) | 0.44 | 0.33 | 0.5 |
| abnormality of higher mental function (HP:0011446) | 0.71 | 0.33 | 0.42 |
| behavioral abnormality (HP:0000708) | 0.46 | 0.33 | 0.37 |
| abnormality of temperature regulation (HP:0004370) | 0.34 | 0.32 | 0.36 |
| abdominal symptom (HP:0011458) | 0.33 | 0.49 | 0.35 |
| **Cluster 2 (653 patient visits, overall inclusivity: 95.56)** | **Exclusion** | **Inclusion** | **IC** |
| abnormal cellular phenotype (HP:0025354) | 0.62 | 0.63 | 0.5 |
| abnormality of the pleura (HP:0002103) | 0.81 | 0.49 | 0.42 |
| nausea and vomiting (HP:0002017) | 0.75 | 0.49 | 0.41 |
| abnormality of digestive system morphology (HP:0025033) | 0.76 | 0.58 | 0.4 |
| functional abnormality of the gastrointestinal tract (HP:0012719) | 0.76 | 0.53 | 0.4 |
| **Cluster 3 (284 patient visits, overall inclusivity: 95.42)** | **Exclusion** | **Inclusion** | **IC** |
| abnormal heart valve morphology (HP:0001654) | 0.68 | 0.36 | 0.5 |
| abnormal cellular phenotype (HP:0025354) | 0.39 | 0.39 | 0.5 |
| abnormal aortic valve physiology (HP:0031652) | 0.66 | 0.37 | 0.43 |
| abnormal heart valve physiology (HP:0031653) | 0.6 | 0.4 | 0.41 |
| arrhythmia (HP:0011675) | 0.5 | 0.37 | 0.39 |
| abnormality of cardiovascular system electrophysiology (HP:0030956) | 0.46 | 0.4 | 0.37 |
| abnormality of the endocrine system (HP:0000818) | 0.43 | 0.4 | 0.37 |
| increased blood pressure (HP:0032263) | 0.37 | 0.5 | 0.37 |
| behavioral abnormality (HP:0000708) | 0.41 | 0.38 | 0.37 |
| abnormal heart morphology (HP:0001627) | 0.5 | 0.46 | 0.37 |

To gain further insight in the differences between the clusters, we projected them onto our derived HPO facets, identifying what percentage of explanatory terms belonged to each facet of HPO (listed in Table 1. These figures can be seen in Table 4. All clusters contain an explanatory term from `abnormal cellular phenotype (HP:0025354)`, although more specific terms than that defining the facet could not be retrieved so as to characterise any of the clusters. In addition, several pairs of clusters also contain explanatory terms from the same facet, such as clusters 1 and 2 containing those from `abnormality of the digestive system`. However, taking into account the overall constitution of facet representations in explanatory sets allows us to see which facets more typify particular clusters. For example, 60% of explanatory terms in cluster 2 are in the `abnormality of the digestive system` facet, while they only account for 20% of those in cluster 1. Together with the high information content and the exclusivity scores for digestive system terms explaining cluster 2, it could be assumed that this cluster identifies admissions for which the primary phenotype is related to the digestive system, with involvement of cellular and respiratory phenotypes as co-morbidities, while cluster 1 is characterised by other phenotypes that may also have involvement of an `abdominal symptom (HP:0011458)`. Most annotations to ICD-9 in the medical ICU in MIMIC are to pumonary and digestive diseases (17.5 and 15.7% respectively).

**Table 4.** Overall facet representation in cluster explanations. Note that percentages may add up to more than one, since ontology terms may belong to several facets. In total, 8 of the 20 HPO facets are represented in the explanations.

| Cluster | Facet | Representation in explanations |
| --- | --- | --- |
| Cluster 1 | abnormality of metabolism/homeostasis | 0.2 |
|  | abnormality of the nervous system | 0.4 |
|  | abnormal cellular phenotype | 0.2 |
|  | abnormality of the digestive system | 0.2 |
| Cluster 2 | abnormal cellular phenotype | 0.2 |
|  | abnormality of the digestive system | 0.6 |
|  | abnormality of the respiratory system | 0.2 |
| Cluster 3 | abnormality of the endocrine system | 0.1 |
|  | abnormality of the cardiovascular system | 0.7 |
|  | abnormality of the nervous system | 0.1 |
|  | abnormal cellular phenotype | 0.1 |
|  | abnormality of blood and bloodforming tissues | 0.1 |

While this gives us some insight into the composition of our set of sampled patient encounters, we can also determine that these clusters do not clearly or uniquely identify individual or groups of phenotypes specific to them. Furthermore, only three clusters were derived from a highly complex and co-morbid dataset. Pearson correlation between the facet representation and the number of annotations in that facet is 0.66, and 0.44 for the relationship between the facet representation and the number of patients the facet represents, both indicating a positive correlation. This indicates that facets with greater numbers of annotations are more likely to appear in explanatory sets. Terms from only 8 of the 20 HPO facets were represented in the overall cluster explanations, although these accounted for 42.44% of all phenotype annotations. It is unclear, however, whether this is because the annotations described by those facets lack underlying structure, are simply uninformative, or whether annotation size bias and more easily recoverable structure is obscuring additional grouping factors. In the next section, we will explore this question by clustering individual facets.

### All Facet Clustering

We calculated one semantic similarity matrix for each facet, by including only annotations from that facet in semantic similarity comparisons of phenotype profiles. We then created clusters for each of these similarity matrices, automatically determining the best number of centers by stepping through numbers of clusters between 3 and 20, selecting the one with the maximal silhouette score. Table 5 presents the results of this analysis, while Figure 6 depicted the visualisations of the clusters with silhouette scores above 0.5.

**Table 5.**
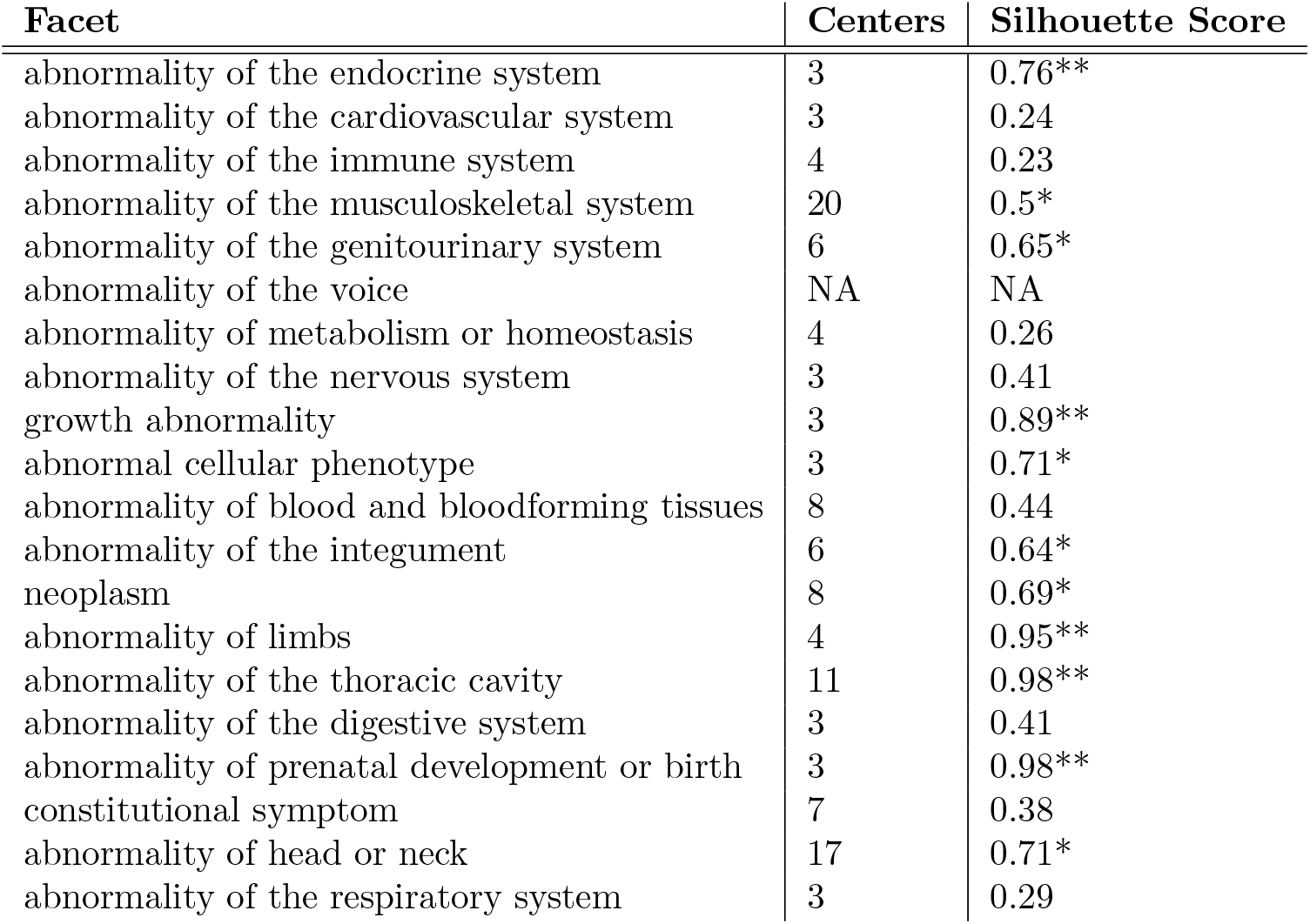
Optimal number of centres and associated silhouette score for each facet. These were obtained by automatically stepping through values of centres between 2 and 20, selecting the one with the greatest silhouette score. Clustering was not performed on the `abnormality of the voice` facet, as there were too few annotations in these faceted profiles. Silhouette scores greater than or equal to 0.5 are marked with a single asterisk, while those greater than 0.75 are marked with two asterisks.

| **Facet** | **Centers** | **Silhouette Score** |
| --- | --- | --- |
| abnormality of the endocrine system | 3 | 0.76** |
| abnormality of the cardiovascular system | 3 | 0.24 |
| abnormality of the immune system | 4 | 0.23 |
| abnormality of the musculoskeletal system | 20 | 0.5* |
| abnormality of the genitourinary system | 6 | 0.65* |
| abnormality of the voice | NA | NA |
| abnormality of metabolism or homeostasis | 4 | 0.26 |
| abnormality of the nervous system | 3 | 0.41 |
| growth abnormality | 3 | 0.89** |
| abnormal cellular phenotype | 3 | 0.71* |
| abnormality of blood and bloodforming tissues | 8 | 0.44 |
| abnormality of the integument | 6 | 0.64* |
| neoplasm | 8 | 0.69* |
| abnormality of limbs | 4 | 0.95** |
| abnormality of the thoracic cavity | 11 | 0.98** |
| abnormality of the digestive system | 3 | 0.41 |
| abnormality of prenatal development or birth | 3 | 0.98** |
| constitutional symptom | 7 | 0.38 |
| abnormality of head or neck | 17 | 0.71* |
| abnormality of the respiratory system | 3 | 0.29 |

**Figure 6.**
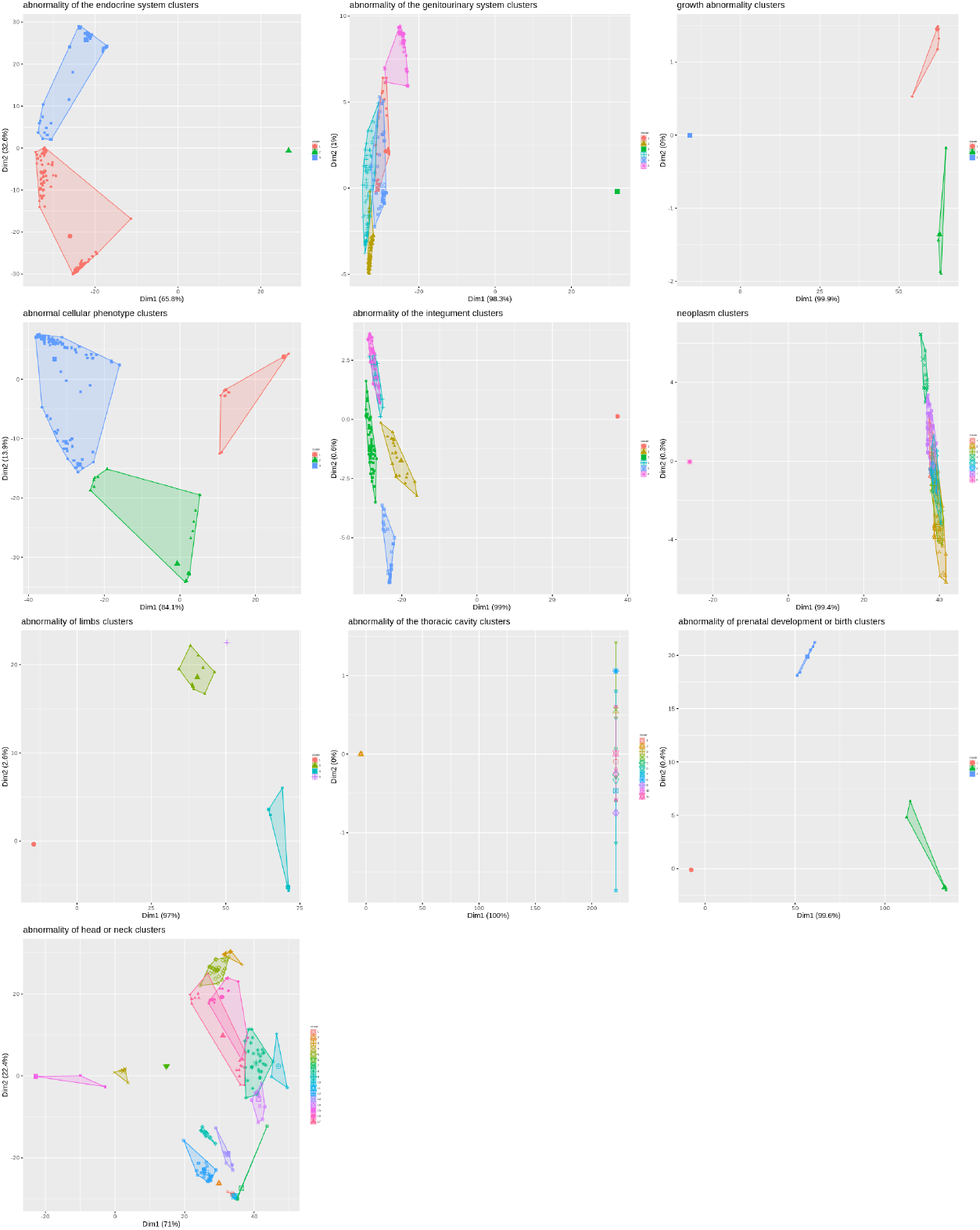
Plots for all facet clusters with a silhouette score above 0.5. In all cases, visualisations show collected relatively close different clusters of patients with relevant keywords, while another cluster contains all patients without relevant keywords.

There was a wide range of highest silhouette scores found amongst faceted clusters, between 0.23 and 0.98. 11 facet clusters had silhouette scores above 0.5, including 5 above 0.75. Pearson correlation between silhouette score and the number of patients was -0.98, while correlation between silhouette score and number of annotations was -0.9, both very strong negative relationships. This

### Cluster Facet Representation in explanations

supports the notion that clustering using facets, reducing the size of phenotype profiles, may lead to higher quality clustering, with more easily recoverable structure.

### Neoplasm Facet Clustering

We decided to further explore the neoplasm facet, since this had a relatively high silhouette score of 0.69, a smaller number of overall clusters, and was not represented in any of the explanations for the overall clustering. The visual representation of the neoplasm facet clusters are presented in Figure 6, while Table 6 lists the explanations derived from these clusters

**Table 6.**
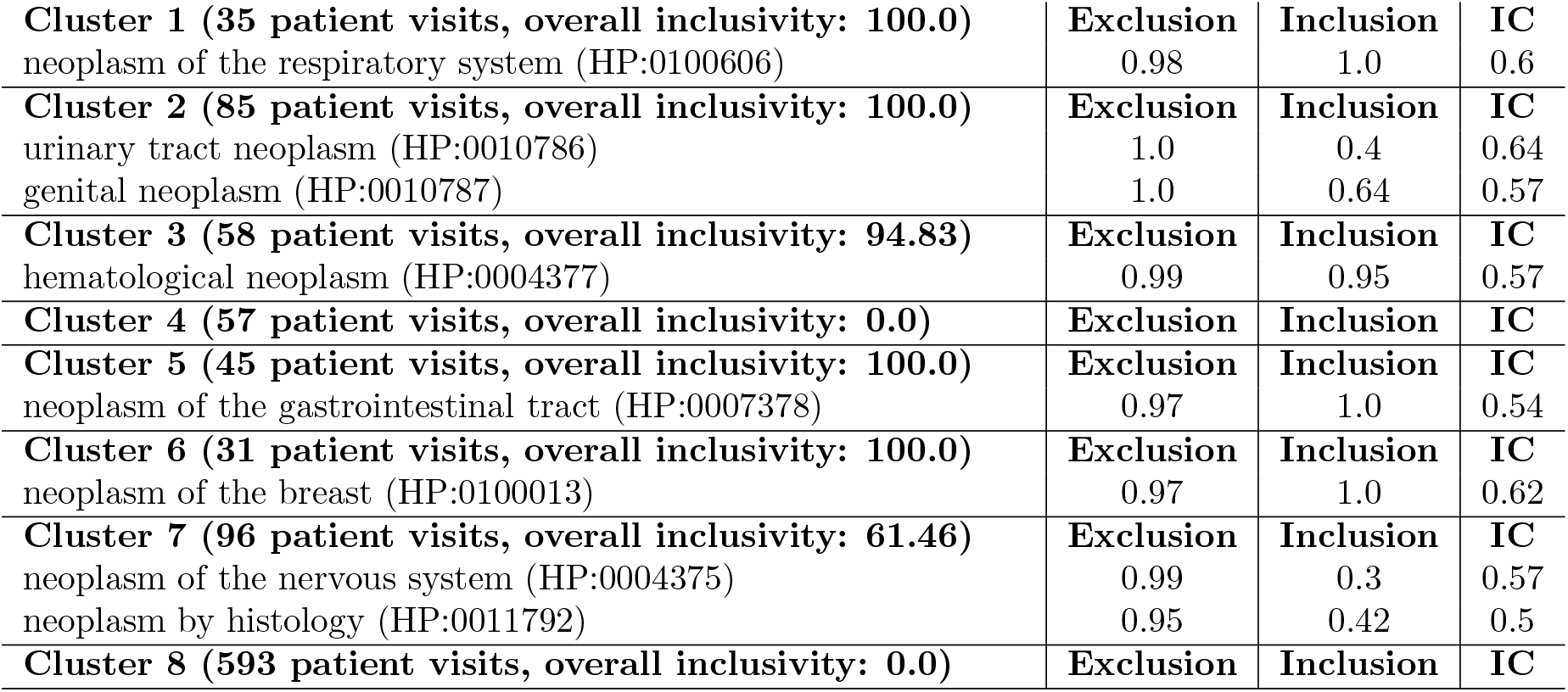
Explanatory sets for clusters in the neoplasm facet.

Extremely clear and distinct explanations were identified for all clusters except 4 and 8. Cluster 8 contains the 593 patients who did not have any neoplastic phenotypes, as shown in Table 1. The 57 patients that appear in cluster 4 did have neoplastic phenotypes, though no characterising groups of phenotypes could be found. It could be that these patients had multiple cancers, or rarer cancers that appeared too infrequently to form a coherent clustering group. Otherwise, all other clusters are strongly inclusively and exclusively characterised by particular groups of cancers.

Nevertheless, patients with neoplasia phenotypes (patients in neoplasm clusters except cluster 8) had a similar distribution in overall clusters as those without neoplasia phenotypes (71.74% in cluster 2 and 19.65% in cluster 3, versus 60.8% in cluster 2 and 34.4% in cluster 3). Assigned percentages in both cases roughly correspond to the distribution of overall patient assignment to those clusters (6.3% in cluster 1, 65.3% in cluster 2, 28.4% in cluster 3), indicating there is no strong association between neoplasia phenotypes and overall cluster assignment, which in turn indicates that neoplasia phenotypes were not widely considered as ‘best match’ terms for the overall semantic similarity comparisons.

The explanation algorithm can be employed to further explore potential co-morbidities relating to neoplasia. In particular, the same descriptions of cluster membership can be used, but the algorithm can be limited to consider only patient phenotype labels in *another* facet. We demonstrated this by applying classes from the `abnormality of the nervous system` and `abnormality of the immune system` facets to the neoplasm cluster memberships. The results are presented in Table 7 and Table 8, respectively. These tables show particular associations between the clusters associated with particular cancer phenotypes and phenotypes from other facets.

**Table 7.**
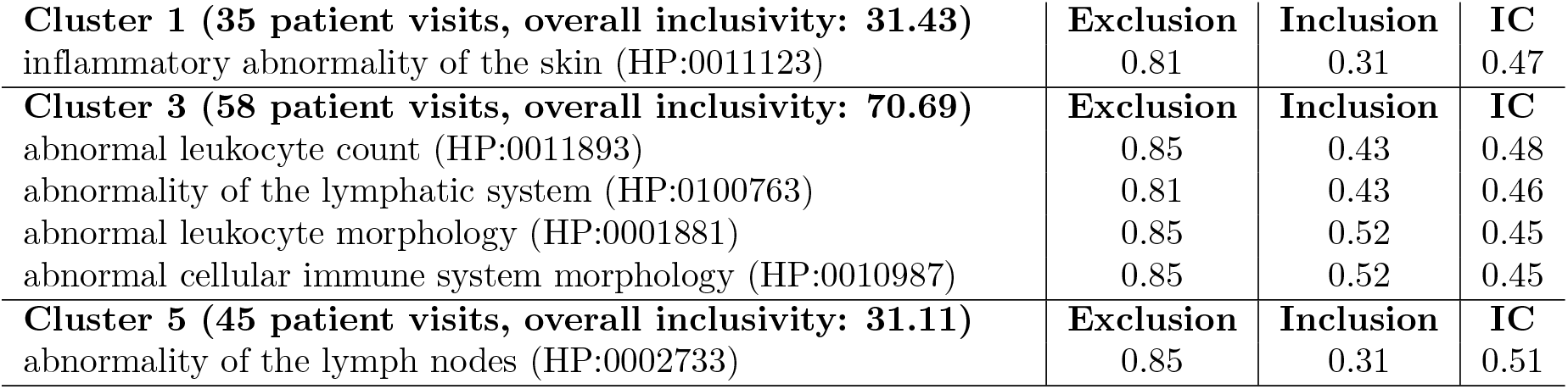
Explanatory terms from `abnormality of the immune system` applied to `neoplasm` facet clusters. Empty explanations not shown (though exclusion scores still include them).

**Table 8.**
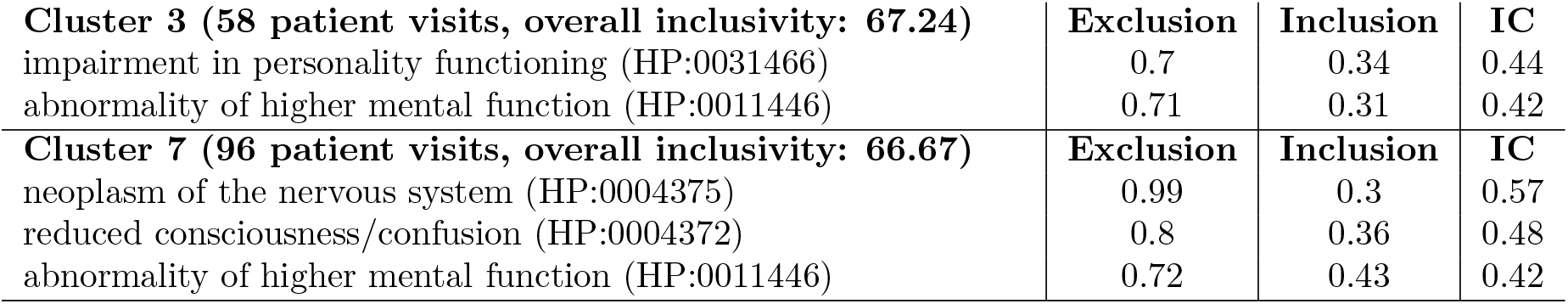
Explanatory terms from `abnormality of the nervous system` applied to `neoplasm` facet clusters. Only non-empty explanations shown (though exclusion scores still include clusters not shown). Used a minimum information content value of 0.4. `neoplasm of the nervous system (HP:0004375)` appears here because it is a member of both the `neoplasm` and the `abnormality of the nervous system facets`.

|  |  |  |  |
| --- | --- | --- | --- |
| **Cluster 3 (58 patient visits, overall inclusivity: 67.24)** | **Exclusion** | **Inclusion** | **IC** |
| impairment in personality functioning (HP:0031466) | 0.7 | 0.34 | 0.44 |
| abnormality of higher mental function (HP:0011446) | 0.71 | 0.31 | 0.42 |
| **Cluster 7 (96 patient visits, overall inclusivity: 66.67)** | **Exclusion** | **Inclusion** | **IC** |
| neoplasm of the nervous system (HP:0004375) | 0.99 | 0.3 | 0.57 |
| reduced consciousness/confusion (HP:0004372) | 0.8 | 0.36 | 0.48 |
| abnormality of higher mental function (HP:0011446) | 0.72 | 0.43 | 0.42 |

Cluster 3 is associated, with hematological neoplasms, including leukemias, lymphomas and non-malignant neoplastic disorders and, as expected, phenotypes characterising these neoplasms are associated with this cluster, namely abnormal leukocyte count, lymphatic system abnormalities and abnormal leukocyte morphology etc. Slightly more unexpected is the association with phenotypes linked to cognitive impairment and compromised higher mental functioning. The segment of this facet that includes these phenotypes is concerned with cognitive function, such as memory, executive function and coordination, rather than affective and personality traits, such as depression. While we know that we are looking at a largely elderly population in MIMIC, the population with this class of cancer would not be expected to show any more specific age-dependent bias in comparison to other common cancers which are also age-associated, such as prostate cancer, and therefore it is unlikely that a comorbid association with dementia and age-related neurodegenerative disease would be generated for this cancer class only. Recently the impact of some hematological neoplasms on higher order functioning has come under scrutiny and whilst there is a well-established impact on these traits. due to radio and chemotherapy [49], it is becoming apparent that cognitive function may also be directly affected as part of the pathology of the disease itself. A recently estimate of impacts on cognitive function in chronic lymphocytic leukemia (CLL) suggests that up to 30% of patients experience significant higher cognitive function impairment [50]. Our method flags a close association in this cluster between patients with hematological malignancies and patients with cognitive impairment, supporting the suggestion of Williams et al. of a more widespread direct interaction with hematological malignancies than previously appreciated.

The association identified between lung malignancies and `inflammatory abnormality of the skin (HP:0011123)` may again reflect therapy-related phenotypes. Drugs targeting the EGF receptor are known to produce skin reactions, erythroderma, eczma-like rashes, dermatitis and paronychia. Erlotinib and gefitinib are widely used to treat small cell (sc) and non-sc lung cancers [51] and these side effects are frequently reported [52].

Cluster 5 associations between gastrointestinal cancers and `abnormality of the lymph nodes (HP:0002733)` likely reflect the common sites of metastasis to abdominal lymph nodes, particularly the mesenteric lymph nodes which are often involved in colonic and rectal adenocarcinomas [53].

Unsurprisingly in cluster 7, an association is found between neoplasms of the nervous system (NS), and abnormalities of higher mental function as well as consciousness and confusion, all well known symptoms of central nervous system tumors. It is however, interesting that while the neoplasia class neoplasm of the nervous system (HP:0004375) includes peripheral NS cancers, the dominant explanatory terms here seem to be associated with the central NS, suggesting that these form a more coherent grouping or are more common in the cohort.

## Discussion

Our work describes the construction of text-derived patient phenotype profiles from clinical narrative text. Using those profiles, we created separate subsets based on facets, which were derived from major subsets of HPO. Table 1 shows that patients are commonly annotated by phenotypes from many different clusters, and that annotation size per-cluster is much smaller than the number of overall annotations per-patient. This indicates that the patient phenotype profiles are highly co- and multi-morbid, with phenotypes across many different biological and categorical systems.

This was confirmed by the overall clustering, which identified only three groups with tendencies towards different kinds of phenotypes, but no clear distinctive groups. These explanations were further dominated by terms from facets with larger numbers of annotations, not including explanatory terms from more than half of the total HPO facets, which nevertheless represented nearly half of all patient phenotypes.

By clustering per-facet, we were able to show that we can identify clusters for faceted subsets of patient phenotype profiles, that are not represented in the overall clusters. This also revealed a strong negative correlation between phenotypic profile size and silhouette score -a trend also exemplified in the poor silhouette score and explanatory terms discerned from the overall clustering experiment. In exploring the neoplasm facet, we found coherent clusters that described different kinds of cancer. We also used explored associations between the neoplasm clusters and other phenotypes, recovering clinically significant relationships, and showing that these were represented in the faceted similarity-based representation of the phenotype profiles. We consider that these results are a proof-of-concept for faceted clustering on a subset of MIMIC-III data, showing that consideration of the semantic similarity of different phenotype facets caters the recovery of groupings and relationships that are not easily recoverable with overall clustering.

In addition, our cluster explanation algorithm allowed us to successfully extract explanatory variables from our clusters. The method also presents a generic approach that could be applied to any dataset that describes entities in terms of both ontology terms, and entity groupings. This is supported by the configurability of the hyperparameters. It may be possible to develop a more intelligent method of determining the optimal trade-off between hyper-parameters. One simple extension could be the automatic suggestion of a default minimum inclusion and exclusion scores on the basis of how many clusters are being explained. There is also the potential that other ways of measuring inclusivity, exclusivity, and specificity could be used -for example, measuring the total number of matching terms across the cluster, instead of the number of members with at least one matching term, or using different information content measures. Exploration of different approaches to MOOPs could also lead to superior solutions.

With respect to clustering, what remains to be done, is to determine a method that enables the re-integration of these scores, clusters, and groupings, into a single representation that minimises loss of information. Such an approach could lead to powerful insights into multi-and co-morbidity, as well as to improve semantic similarity based classification using text-derived phenotypes, such as that described by our previous work [11]. One method of approaching this specific to clustering problems could be the consideration of Multi-View Clustering, which could consider each measure of facet-wise similarity as a different view of the same patient admission [54], which could then be further analysed to determine an optimal set of clusters.

In our work, we chose direct subclasses of `Phenotypic abnormality (HP:0000118)` for our facets, because these split the phenotypes appropriately by recognisable means (i.e. biological system). These should be a suitable starting point for any similar experiment. The approach, however, could easily be applied to many ontologies, by selecting a reasonable high-level class with which to generate facet categories. For example, the Disease Ontology (DO) [55] defines eight direct subclasses of `disease (DOID:4)`, including `disease by infectious agent (DOID:0050117)` and disease of mental health (DOID:150). The choice of a facet superclass does not necessarily have to be a high-level class. For domain or context-specific investigations, facets can be drawn from more specific classes, such as `abnormal heart morphology (HP:0001627)`, whose direct subclasses describe many different kinds of abnormal heart morphology, which are further explicated by transitive subclasses. In addition, using Komenti enables for facets to be drawn from the subclasses of complex class descriptions, even across multiple ontologies, such as `‘part of’ some apoptosis`.

However, while manually chosen facets provide different sets of features by which to explore entities through semantic similarity, the potential remains for information to be lost within a facet in the same way as through calculating similarity as an entire ontology. For example, there may be several potential grouping factors within the cardiac facet. To minimise this effect, an approach could be developed to automatically select facets based on different hotspots of high corpus-based information content in an ontology. This would ensure that different areas of importance in the ontology are captured by the chosen set of facets. We also noted, in accordance with our hypotheses, that silhouette score has a strong negative correlation with annotation size, and it could be that optimising facet groups, based on a maximum profile size to limit performance issues, could be employed.

One of the limitations of this approach is that it involved generating many different clusters, from many different facets, without individual attention given to configuring parameters that may have led to an improved representation. Optimisation of silhouette score was automated, by stepping through number of centres. However, we did not consider different values for creation of the optimisation, kmeans algorithm, or semantic similarity measure. Depending on any underlying structure of facet similarity matrices, these choices could make the difference between successful or unsuccessful clustering. It’s possible that different facets contain different underlying structure, which require different settings and algorithms to evaluate. We expect that a further study, exploring further different hyper-parameters, measures, and options for text-derived semantic similarity clustering, may lead to reasonable defaults. For example, different measures of cluster evaluation could lead to better identification of optimal parameters, such as the gap statistic.

In terms of the application of the explanation algorithm to find explanations for facet clusters from another cluster, results were skewed by the polyhierarchy of HPO. Facets do not necessarily uniquely belong to a single cluster. This is evidenced by the `abnormality of the nervous system` explanations for the `neoplasm` clusters, which included `neoplasm of the nervous system (HP:0004375)` as an explanation for the neoplasm cluster already associated with `neoplasm of the nervous system (HP:0004375)`. This is not an error since the phenotype is a subclass of both facets, however it strongly skews the explanations towards any classes that are members of the facet by which the admissions were clustered, possibly to the exclusion of other classes. It may be possible to explore the exclusion of candidate explanatory classes from the ‘native’ facet through provision to the algorithm of another parameter identifying it.

Another limitation of the study was the constraint of admission selection to those with primary diagnoses mapped to DO. In our results, we showed that in one facet with poor annotation representation `abnormality of prenatal development or birth`, that few relevant ICD-9 codes were mapped to DO. As such, we can expect that facet representation was skewed against patients with fewer DO mappings. However, for other facets, representation was in line with expected proportions of admission diagnoses in MIMIC-III. The goal of this investigation, however, is not so much to produce the most realistic representation of MIMIC-III admissions overall, but to produce a better representation of the patients we *did* select. To gain a truer representation of phenotype representation across MIMIC-III, the experiment could be performed again without constraining by mapped diagnosis, although this would limit secondary analysis for diagnosis classification.

Another cause for concern with text-derived phenotypes is their noisiness. In our case, the phenotype profile is simply the set of annotations made for that admission, or the set of ontology terms recognised in their clinical narrative. However, the appearance of a phenotype in text does not necessarily imply that the patient has that phenotype. To some extent, this is mitigated by corpus-based measures of information content, however previous work has shown that curated text phenotypes performed better for the task of predicting rare diseases [12]. This implies that in addition to splitting semantic similarity calculations into different facets with smaller annotation size, methods for automated, semi-automated, or manual curation of text-derived phentoypes could also lead to improved performance. One method of doing this could be to create a modified measure of information content, that measures entity coverage instead of total annotation frequency in corpus. By doing this, phenotypes that appear only a small number of times per patient, but are represented in most patients, would be more correctly scaled.

## Conclusions

Our work describes the successful categorisation of text-derived patient phenotype profiles into facets derived from major areas of the Human Phenotype Ontology (HPO). In doing this, we showed that in our setting, phenotypic expression falls widely across different areas of HPO. Using these faceted phenotype profiles, we successfully developed and implemented a semantic clustering approach, comparing overall clustering with faceted clustering. To evaluate these clusters, we developed and presented a novel algorithm for characterising ontology-based groupings. Using this method, we showed that, in our setting, faceted semantic clustering provides clinically meaningful insights into text-derived patient phenotype profiles.

## Data Availability

Patient data is available via MIMIC. Software is available by the attached link.

https://github.com/reality/facetsim

## Ethical approval

This work makes use of the MIMIC-III dataset, which was approved for construction, de-identification, and sharing by the BIDMC and MIT institutional review boards (IRBs). Further details on MIMIC-III ethics are available from its original publication (DOI:10.1038/sdata.2016.35). Work was undertaken in accordance with the MIMIC-III guidelines.

## Availability of data and material

Komenti is an open source text mining framework, and is available under an open source licence from https://github.com/reality/komenti. The code used to sample MIMIC, and run similarity experiments is available from https://github.com/reality/miesim/. The annotated MIMIC dataset is not made publicly available, because researchers are required to meet ethical conditions to access MIMIC-derived datasets. To access this dataset, please contact the corresponding author directly.

## Competing interests

The authors declare that they have no competing interests.

## Acknowledgements

GVG and LTS acknowledge support from support from the NIHR Birmingham ECMC, NIHR Birmingham SRMRC, Nanocommons H2020-EU (731032) and the NIHR Birmingham Biomedical Research Centre and the MRC HDR UK (HDRUK/CFC/01), an initiative funded by UK Research and Innovation, Department of Health and Social Care (England) and the devolved administrations, and leading medical research charities. The views expressed in this publication are those of the authors and not necessarily those of the NHS, the National Institute for Health Research, the Medical Research Council or the Department of Health. RH, PNS and GVG were supported by funding from King Abdullah University of Science and Technology (KAUST) Office of Sponsored Research (OSR) under Award No. URF/1/3790-01-01. AK was supported by by the Medical Research Council (MR/S003991/1) and the MRC HDR UK (HDRUK/CFC/01). PNS and GVG acknowledge the support of the Alan Turing Institute, UK.

## Appendix

**Table 9.** Parameters used for explanation algorithm. Parameters are defined in Algorithm 1.

| Name | Value |
| --- | --- |
| MAX_IC | 0.8 |
| MIN_IC | 0.4 |
| MAX_INCLUSION | 0.95 |
| MIN_INCLUSION | 0.3 |
| MAX_EXCLUSION | 0.95 |
| MIN_EXCLUSION | 0.3 |
| MAX_TOTAL_INCLUSION | 0.95 |
| STEP | 0.05 |

## Notes

### Competing Interest Statement

The authors have declared no competing interest.

